# What Patients Reward and Punish in Physician Communication: A Framework-Based Analysis of 62,319 Indian Reviews

**DOI:** 10.64898/2026.07.26.26358948

**Authors:** Rajshri Mallabadi, Shweta Sharma, Bidita Khandelwal, Meenal Mohgaonkar, Venkatarao Epari

## Abstract

**Background:** Patient-generated online reviews contain detailed accounts of physician communication that remain systematically underused in health services research. Prior computational analyses have either applied service quality frameworks without grounding in clinical communication theory, or have been geographically and specialty-restricted.

**Objective:** To examine which physician communication dimensions predict patient recommendation, switching intent, and retention across specialties and cities in India.

**Methods:** We analyzed 62,319 patient reviews from the Practo platform (10 cities, 8 specialties) coded against established clinical communication frameworks, the Kalamazoo Consensus Statement, Calgary-Cambridge Guide, and SPIKES protocol, adapted into a 12-dimension codebook. Expert confirmatory review on 200 stratified reviews yielded 98.5% agreement between reviewer and model labels across 11 communication dimensions. Binary recommendation was designated the primary outcome; recommendation intensity is reported as secondary. Logistic regression with 1,000 bootstrap iterations examined associations between communication dimensions and recommendation and switching intent, adjusting for treatment outcome, cost, wait time, and specialty.

**Results:** Reassurance (OR 2.07, 95% CI 1.86–2.42) and empathy (OR 1.89, 95% CI 1.79–2.00) were the strongest positive predictors of recommendation. Excessive directiveness (OR 0.62, 95% CI 0.60–0.64) and rushedness (OR 0.65, 95% CI 0.64–0.67) were the dominant negative predictors; directiveness was present in 33.3% of switching-intent reviews. Cost asymmetry was pronounced: cost concerns reduced recommendation rates by 54.2 percentage points; positive cost comments increased recommendation by 6.2 points. Specialty variance in recommendation intensity exceeded city variance (ratio 2.63).

**Conclusions:** Excessive directiveness and rushedness were the communication behaviors most strongly associated with adverse behavioral outcomes in this corpus; reassurance and empathy are the strongest drivers of recommendation. These effects are modified by specialty context in ways that align with Kalamazoo, Calgary-Cambridge, and SPIKES theoretical predictions. The specialty-stratified findings provide a direct empirical basis for competency-based communication curricula in undergraduate and postgraduate medical education.

## Introduction

Patient satisfaction research has historically relied on administered surveys, such as Press Ganey or the Consumer Assessment of Healthcare Providers and Systems (CAHPS), which are constrained by structured response formats, sampling biases, and social desirability effects. Online physician review platforms bypass these limitations. As Hong et al. documented in their systematic review, patient online reviews have grown substantially in volume and influence on physician selection ^[1]^, capturing dimensions of care that structured instruments frequently miss.

The Practo platform, India’s largest physician booking and review service, has accumulated millions of such reviews. Prior analysis by Dhakate and Joshi examined 38,019 reviews from Delhi and Bangalore using inductive sentiment analysis ^[2]^, demonstrating the corpus’s analytical potential but remaining geographically restricted and without clinical communication framework anchoring.

Three frameworks define the evidence base for effective physician communication. The Kalamazoo Consensus Statement identified seven essential communication tasks ^[3]^. The Calgary-Cambridge Guide operationalized these into a five-stage structured interview ^[4]^. The SPIKES protocol addressed breaking bad news in oncology and serious illness contexts ^[5]^. These three frameworks are complementary rather than overlapping: Kalamazoo defines the essential tasks, Calgary-Cambridge provides the process structure, and SPIKES addresses the affective and prognostic dimensions that the other two do not cover.

Zhang et al. applied the HSQ-5D framework, derived from Donabedian and SERVQUAL, to Chinese platform reviews ^[6]^, differentiating their work by context and analytical framework. The physician review literature has noted persistent data quality concerns ^[7]^, and most published analyses have been conducted in Western contexts ^[1,8]^.

To our knowledge, no published study has operationalized the Kalamazoo Consensus Statement, the Calgary-Cambridge Guide, and the SPIKES protocol as a unified coding framework for large-scale analysis of patient-generated physician reviews. The present study does so, examining 62,319 reviews across all five Indian geographic regions, 10 cities, and 8 specialties, with pre-specified hypotheses about which communication dimensions predict recommendation, switching intent, and retention.

This study introduces a framework for translating established clinical communication theory into computationally measurable constructs that can be applied to large-scale patient-generated narratives. The resulting analyses provide empirical evidence on the communication behaviors associated with patient recommendation, switching intent, and retention in the Indian context, while illustrating the feasibility of implementing theory-driven communication coding at scale through LLM-assisted annotation and expert confirmatory review.

## Methods

### Study Design and Ethical Considerations

This is an observational computational study using publicly available patient review data. Reviews were collected from publicly accessible physician profile pages on the Practo platform (practo.com). No login, authentication, subscription, or bypass of technical access controls was required to access the review content at the time of collection. Doctor names were replaced with platform-assigned slugs. No identifiable patient information was used; review content was publicly visible, and physician identities were pseudonymized, aligning with prevailing norms for digital-trace research. The study analyzed existing publicly available content without interacting with users or altering platform functionality. This study was not pre-registered. Hypotheses were developed a priori from the clinical communication frameworks and refined following a rule-based pilot on the full corpus prior to LLM-assisted annotation.

### Data Source and Corpus Construction

Reviews were collected from Practo.com between March and April 2026, targeting physician profiles across 10 cities spanning all five Indian geographic regions (Delhi, Mumbai, Bangalore, Kolkata, Chennai, Hyderabad, Pune, Ahmedabad, Jaipur, Indore) and 8 specialties (General Medicine, General Surgery, Cardiology, Dermatology, Psychiatry, Endocrinology, Diabetology, Oncology). The corpus was frozen on 20 April 2026 at 62,319 reviews across 80 specialty-city cells. After excluding invalid or non-patient reviews (n = 920) and excessively short reviews (n = 223), the analyzable corpus comprised 61,176 reviews.

### Theoretical Framework and Codebook

The analytical codebook was developed from three complementary clinical communication frameworks: Kalamazoo ^[3]^, Calgary-Cambridge ^[4]^, and SPIKES ^[5]^. The final codebook comprised 12 binary communication dimensions, 7 nuisance controls, and 5 outcome-tier variables. The enablement dimension was tightened to require explicit evidence of skill or knowledge transfer rather than vague encouragement. Full definitions and decision rules are available from the authors on request.

### LLM-Assisted Coding and Outcome Designation

LLM-assisted annotation has been shown to match or exceed crowdsourced annotation reliability on a range of text classification tasks ^[9]^. The coding protocol was piloted on 200 reviews using Claude Sonnet (Anthropic, 2025) ^[10]^, then applied to the full corpus using GPT-4.1-mini (OpenAI, 2025) ^[11]^ via the OpenAI Batch API. The codebook and eight few-shot examples were supplied as the system message (~4,350 tokens); each review was supplied as the user message. Temperature was set to 0; output tokens were capped at 600 per review. Total annotation cost was approximately USD 100.

Manual annotation of more than 61,000 reviews across multiple communication dimensions would have required substantial expert annotation effort. LLM-assisted annotation therefore served as a scalable implementation of a predefined theory-driven codebook rather than as a mechanism for hypothesis generation. Inter-model calibration confirmed stable agreement on core communication dimensions (kappa range 0.79–1.00). A systematic scale compression of approximately 0.55 points was observed for recommendation intensity. Accordingly, binary recommendation (the Practo structured YES/NO field) is designated the primary outcome for all hypothesis tests. Recommendation intensity is retained as a secondary outcome.

### Expert Confirmatory Review

A stratified random sample of 200 reviews (25 per specialty) was reviewed by the lead author (RM), a clinically trained researcher familiar with the codebook. GPT-4.1-mini predictions were presented as a starting point; the reviewer confirmed or overrode each label independently. This procedure constitutes expert confirmatory review of model output rather than fully independent double-coding, and is disclosed as such.

Expert confirmatory agreement was 98.5% across 11 of the 12 binary communication dimensions, with 22 labels changed across 200 reviews. The twelfth dimension, stigma blame, had zero positive cases in the confirmatory sample, rendering kappa computationally undefined; it is excluded from the agreement table but retained in the analytical dataset. Agreement on recommendation intensity was 99.5% with a mean absolute difference of 0.02 scale points. Label-level statistics are reported in Supplementary Table A1.

### Outcome Variables

Four outcome variables were examined: binary recommendation (Practo structured YES/NO field, primary); recommendation intensity (0–3 ordinal, secondary); switching intent (binary, coded from non-return language); retention signal (binary, coded from repeat visit evidence).

### Statistical Analysis

Primary hypothesis testing used logistic regression with binary recommendation and switching intent as outcomes. All 12 communication dimensions, nuisance controls, and specialty fixed effects were included as predictors; all predictors were standardized. ORs and 95% CIs were estimated using 1,000 bootstrap iterations with stratified sampling. Specialty versus city variance in recommendation intensity was tested by computing the variance of group means. Analyses were conducted in Python 3.14 using scikit-learn 1.5. Regression analyses used complete-case observations, resulting in an analytic sample of 60,526 reviews; records with missing outcome or predictor values were excluded. City was treated as a descriptive stratifying variable rather than a multilevel predictor; the primary variance decomposition showed specialty effects dominated city effects at a ratio of 2.63, justifying this decision.

Sensitivity analyses excluding the highest-volume city (Bangalore, 19.3% of corpus) and the smallest specialty group (oncology, n=1,803) confirmed that key odds ratios for reassurance, empathy, excessive directiveness, and rushedness remained stable within ±0.05 of the primary estimates. The codebook, annotation pipeline, and statistical analysis scripts are publicly available at https://doi.org/10.17605/OSF.IO/UARJW

### Plain-language summary of methods

For readers unfamiliar with computational methods: this study collected 62,319 written reviews that patients had posted online about their doctors. An artificial intelligence system, trained using established clinical communication guidelines, read each review and identified whether it mentioned specific communication behaviors, such as whether the doctor listened carefully, explained things clearly, or seemed rushed. A senior clinician then reviewed a stratified sample of 200 reviews, confirming or overriding the model labels; overall agreement was 98.5%.

Statistical analysis identified which communication behaviors appeared most often in reviews where patients said they would recommend the doctor, and which appeared most often in reviews where patients said they would not return or would warn others away. The results therefore reflect patterns across more than 61,000 real patient experiences across India.

## Results

### Corpus Description

Of 62,319 reviews, 61,176 (98.2%) were analyzable. Descriptive analyses use this full analyzable corpus; regression analyses are restricted to 60,526 complete-case observations as described in Section 2.7. Recommendation balance was 93.8% YES and 6.3% NO, consistent with the positivity bias documented for online physician review platforms. Tables 1a and 1b report the distribution by specialty and city respectively.

**Table 1a.** Corpus distribution by specialty.

| Specialty | n | % |
| --- | --- | --- |
| General physician | 12,395 | 20.3% |
| Dermatologist | 11,600 | 19.0% |
| General surgeon | 10,210 | 16.7% |
| Psychiatrist | 9,264 | 15.1% |
| Cardiologist | 6,623 | 10.8% |
| Endocrinologist | 4,664 | 7.6% |
| Diabetologist | 4,617 | 7.5% |
| Oncologist | 1,803 | 2.9% |
| Total | 61,176 | 100% |

**Table 1b.** Corpus distribution by city.

| City | n | % |
| --- | --- | --- |
| Bangalore | 11,849 | 19.4% |
| Delhi | 9,321 | 15.2% |
| Mumbai | 8,175 | 13.4% |
| Hyderabad | 7,957 | 13.0% |
| Pune | 7,863 | 12.9% |
| Chennai | 6,629 | 10.8% |
| Jaipur | 2,848 | 4.7% |
| Kolkata | 2,805 | 4.6% |
| Indore | 1,924 | 3.1% |
| Ahmedabad | 1,805 | 2.9% |
| Total | 61,176 | 100% |

### Communication Dimension Prevalence

Table 2 reports the prevalence of each communication dimension. High-prevalence dimensions (empathy 45.8%, explanation clarity 35.1%, listening 31.6%, reassurance 18.2%) reflect dimensions patients mention frequently in unstructured text. Low-prevalence dimensions, notably excessive directiveness (1.6%) and rushedness (1.9%), carry disproportionate behavioral consequences.

**Table 2.** Communication dimension prevalence (n = 61,176)

| Dimension | n | % | Dimension | n | % |
| --- | --- | --- | --- | --- | --- |
| Empathy and concern | 27,692 | 45.8% | Cost transparency | 727 | 1.2% |
| Explanation clarity | 21,239 | 35.1% | Excessive directiveness | 953 | 1.6% |
| Listening | 19,133 | 31.6% | Rushedness | 1,161 | 1.9% |
| Reassurance | 11,013 | 18.2% | Shared decision-making | 1,709 | 2.8% |
| Staff conduct | 6,746 | 11.1% | Stigma/blame | 45 | 0.1% |
| Retention signal | 6,382 | 10.5% | Switching intent | 1,171 | 1.9% |
| Respect (general) | 5,364 | 8.9% | Anti-recommendation | 529 | 0.9% |
| Enablement | 3,879 | 6.4% | Cost expensive | 1,206 | 2.0% |

### H1: Communication Dimensions and Recommendation

In adjusted logistic regression (Table 3), reassurance (OR 2.07, 95% CI 1.86–2.42) and empathy (OR 1.89, 95% CI 1.79–2.00) were the strongest positive communication predictors of binary recommendation after treatment outcome (OR 4.33, 95% CI 3.79–5.08). Enablement (OR 1.52, 95% CI 1.35–1.71) and shared decision-making (OR 1.38, 95% CI 1.22–1.55) were also significant positive predictors. Figure 2 illustrates the forest plot.

**Table 3.** Logistic regression: binary recommendation (YES=1), n = 60,526.

| Predictor | OR | 95% CI (1,000 bootstrap) | Sig. |
| --- | --- | --- | --- |
| Treatment outcome (positive) | 4.33 | [3.79, 5.08] | * |
| Reassurance and validation | 2.07 | [1.86, 2.42] | * |
| Empathy and concern | 1.89 | [1.79, 2.00] | * |
| Enablement and self-management | 1.52 | [1.35, 1.71] | * |
| Cost reasonable | 1.52 | [1.36, 1.83] | * |
| Shared decision-making | 1.38 | [1.22, 1.55] | * |
| Wait acceptable | 1.25 | [1.15, 1.38] | * |
| Explanation clarity | 1.17 | [1.12, 1.23] | * |
| Staff conduct | 1.04 | [1.00, 1.09] | * |
| Listening attentiveness | 1.02 | [0.97, 1.07] | ns |
| Cost transparency | 0.89 | [0.86, 0.92] | * |
| Respect (general) | 0.85 | [0.81, 0.88] | * |
| Excessive directiveness | 0.62 | [0.60, 0.64] | * |
| Rushedness | 0.65 | [0.64, 0.67] | * |
| Cost expensive | 0.70 | [0.68, 0.72] | * |
| Wait excessive | 0.67 | [0.66, 0.69] | * |
| Treatment outcome (negative) | 0.60 | [0.59, 0.62] | * |

\* 95% CI excludes 1.0 (1,000 bootstrap iterations, stratified sampling); ns = not significant. All predictors standardized. General physician = reference specialty. Two adjusted ORs warrant brief explanation. Respect (general) (OR 0.85) is coded when patients explicitly mention interpersonal dignity in either direction — positive or negative. In the adjusted model, after controlling for dimensions that dominate positive consultations, the residual signal tilts toward negative mentions, producing an OR below 1.0. Cost transparency (OR 0.89) follows the same logic: negative instances (hidden charges, surprise billing) slightly outnumber positive ones. Both dimensions function more as markers of noteworthy encounters than as unidirectional quality signals.

**Figure 1.**
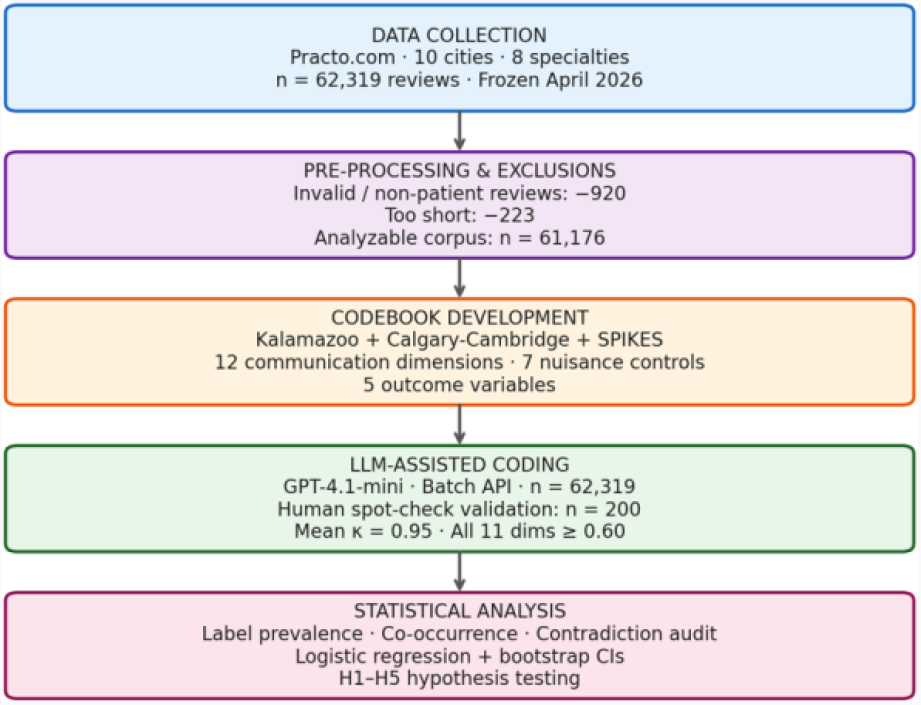
Study design, data selection, and analytical workflow.

**Figure 2.**
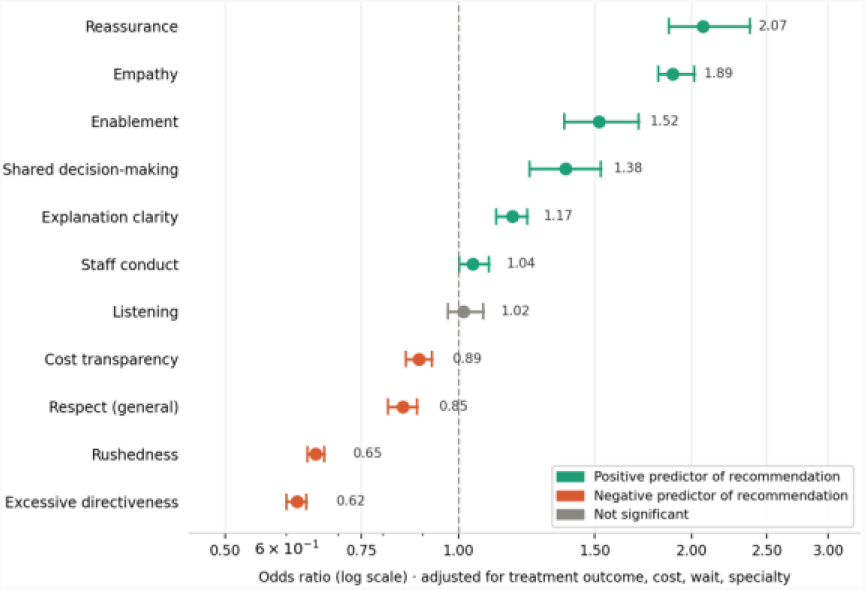
Communication predictors of binary recommendation. Logistic regression ORs with 95% bootstrap CIs, n = 60,526. * CI excludes 1.0.

Excessive directiveness (OR 0.62, 95% CI 0.60–0.64) and rushedness (OR 0.65, 95% CI 0.64– 0.67) were the dominant negative predictors. In unadjusted analysis, reviews mentioning excessive directiveness showed a recommendation rate of 22.9% versus 94.9% in reviews without this dimension — a 72 percentage-point gap. The equivalent gap for rushedness was 66 percentage points. These absolute rate differences substantially exceed the positive communication effects, consistent with the asymmetric impact of negative versus positive experiences documented in behavioral research.

### H2: Cost Asymmetry

Reviews mentioning high or unfair costs (n = 1,206) showed a recommendation rate 54.2 percentage points lower than reviews without this content (38.7% vs. 92.9%). Reviews mentioning reasonable fees (n = 1,521) showed only a 6.2 percentage-point increase (99.3% vs. 93.1%). The negative-to-positive cost effect ratio was approximately 9:1. In the switching intent model, cost_expensive was a significant positive predictor of switching (OR 1.20, 95% CI 1.16– 1.25).

### H3: Specialty-Modified Communication Effects

Specialty context substantially modified which communication dimensions were most salient. Reassurance dominated in oncology (31.4% prevalence) and psychiatry (29.3%), consistent with SPIKES protocol priorities and the centrality of the therapeutic alliance in psychiatric practice. In general surgery, reassurance again led (24.4% prevalence), consistent with perioperative anxiety being a primary patient concern. Enablement was highest in endocrinology and diabetology (both 9.8%), consistent with chronic disease self-management demands. Staff conduct was highest in dermatology (17.3%), reflecting the chain-clinic structure common to Indian dermatology practice. Figure 3 shows the full prevalence heatmap by specialty.

**Figure 3.**
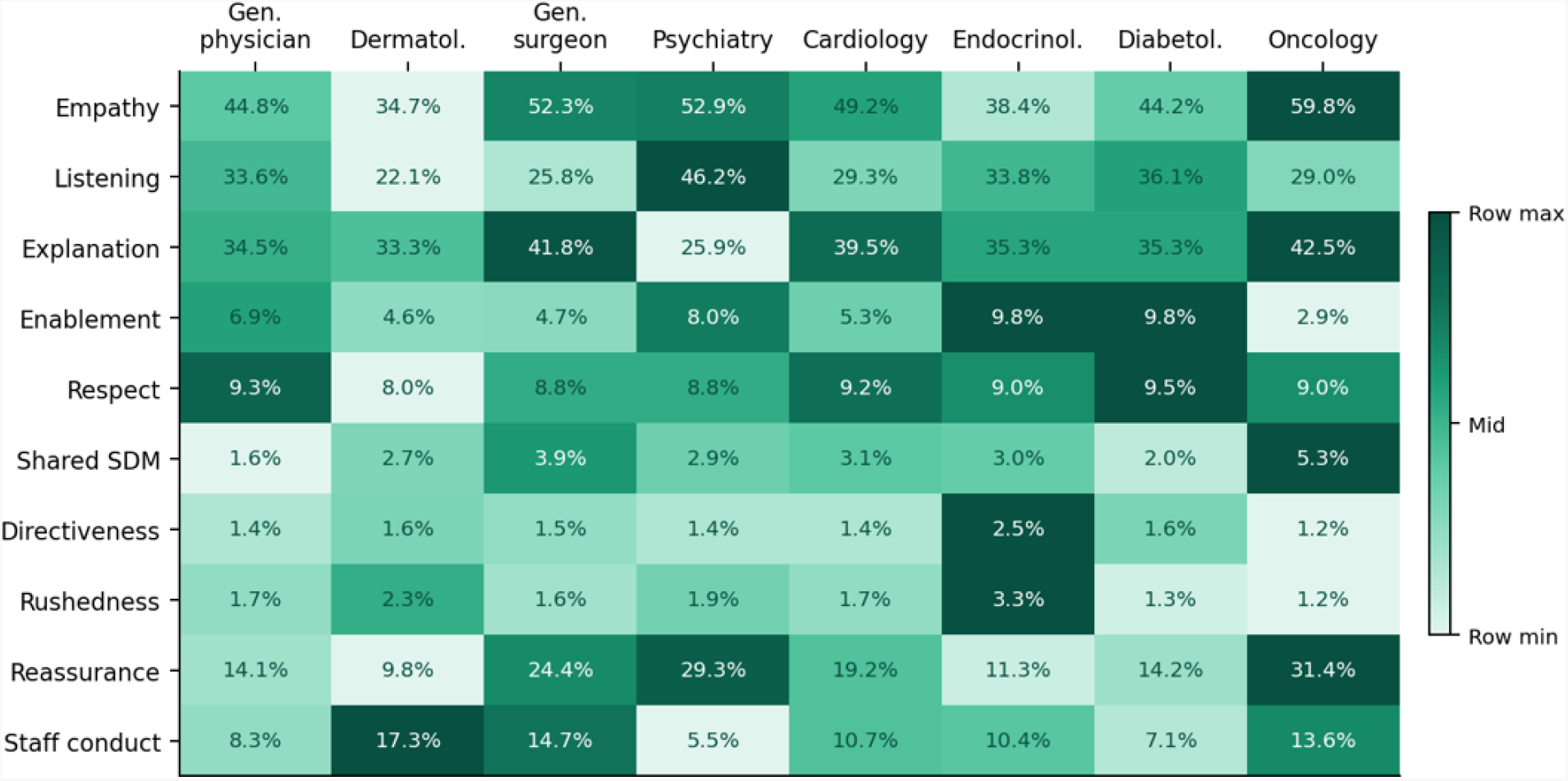
Communication dimension prevalence by specialty (%). Each cell shows % of reviews in that specialty mentioning the dimension. Color scale normalized within each row to highlight cross-specialty variation.

Table 4 reports the top three predictors of recommendation intensity within each specialty. Empathy appeared in the top three across every specialty, while the relative weight of reassurance, shared decision-making, and explanation clarity varied by clinical context.

**Table 4.** Top communication predictors of recommendation intensity by specialty.

| Specialty | Largest intensity differential | 2nd largest differential | 3rd largest differential |
| --- | --- | --- | --- |
| General physician | Empathy (+0.36) | Shared SDM (+0.36) | Reassurance (+0.31) |
| Dermatologist | Directiveness (+0.36) † | Shared SDM (+0.25) | Empathy (+0.22) |
| General surgeon | Reassurance (+0.40) | Empathy (+0.37) | Shared SDM (+0.34) |
| Psychiatrist | Directiveness (+0.45)† | Empathy (+0.32) | Reassurance (+0.31) |
| Cardiologist | Shared SDM (+0.42) | Empathy (+0.32) | Directiveness (+0.27)† |
| Endocrinologist | Directiveness (+0.74)† | Empathy (+0.32) | Respect (+0.24) |
| Diabetologist | Directiveness (+0.41)† | Empathy (+0.27) | Reassurance (+0.22) |
| Oncologist | Directiveness (+0.68)† | Empathy (+0.47) | Reassurance (+0.46) |

† The positive intensity differential for directiveness reflects bidirectional intensity coding, explained in Section 4.2. In the binary recommendation model, excessive directiveness is the strongest negative predictor across all specialties (OR range 0.54–0.71).

A note on the directiveness intensity differential: recommendation intensity (0–3) was coded to capture signal strength, not valence; intensity 3 is assigned to emphatic reviews in both positive and negative directions. Reviews mentioning excessive directiveness are disproportionately emphatic regardless of valence, which inflates their mean intensity score. The binary recommendation model shows the true direction: excessive directiveness is consistently the strongest negative predictor across all specialties. Readers should not interpret the intensity pattern in Table 4 as evidence of clinical benefit.

### H4: Specialty Versus City Variance

When we compared how much recommendation scores varied by specialty versus by city, specialty explained more than twice as much of the variation (specialty variance 0.01035; city variance 0.00393; ratio 2.63). In practical terms, whether a patient consulted a psychiatrist versus an oncologist mattered more to their experience than whether they were in Delhi versus Mumbai. Oncology and general surgery showed the highest specialty means (1.60 and 1.57); general physician and endocrinology showed the lowest (1.33 each). This pattern is consistent with clinical-context factors — nature of illness, prognosis, and relationship duration, shaping communication expectations more strongly than regional or geographic factors.

### H5: Communication Content and Review Engagement

Mean communication dimension count increased monotonically with recommendation intensity: reviews at intensity 0 mentioned 1.19 dimensions on average; intensity 1, 1.37; intensity 2, 1.96; intensity 3, 2.10. More emotionally engaged reviews contain richer communication content, supporting a reframing of this hypothesis around engagement rather than valence.

### Switching Intent and Retention

Switching intent was present in 1,171 reviews (1.9%). Communication behaviors were associated with expressions of switching intent in a consistent pattern: excessive directiveness was present in 33.3% of switching-intent reviews versus 1.4% of non-switching reviews; rushedness appeared in 24.3% versus 1.5%. Reassurance appeared in only 0.07% of switching-intent reviews, making it the strongest protective factor. In adjusted logistic regression (Table 5), excessive directiveness (OR 1.34, 95% CI 1.30–1.38) and rushedness (OR 1.18, 95% CI 1.14– 1.22) showed the strongest positive associations with switching intent. Figure 4 shows switching intent rates by dimension.

**Table 5.**
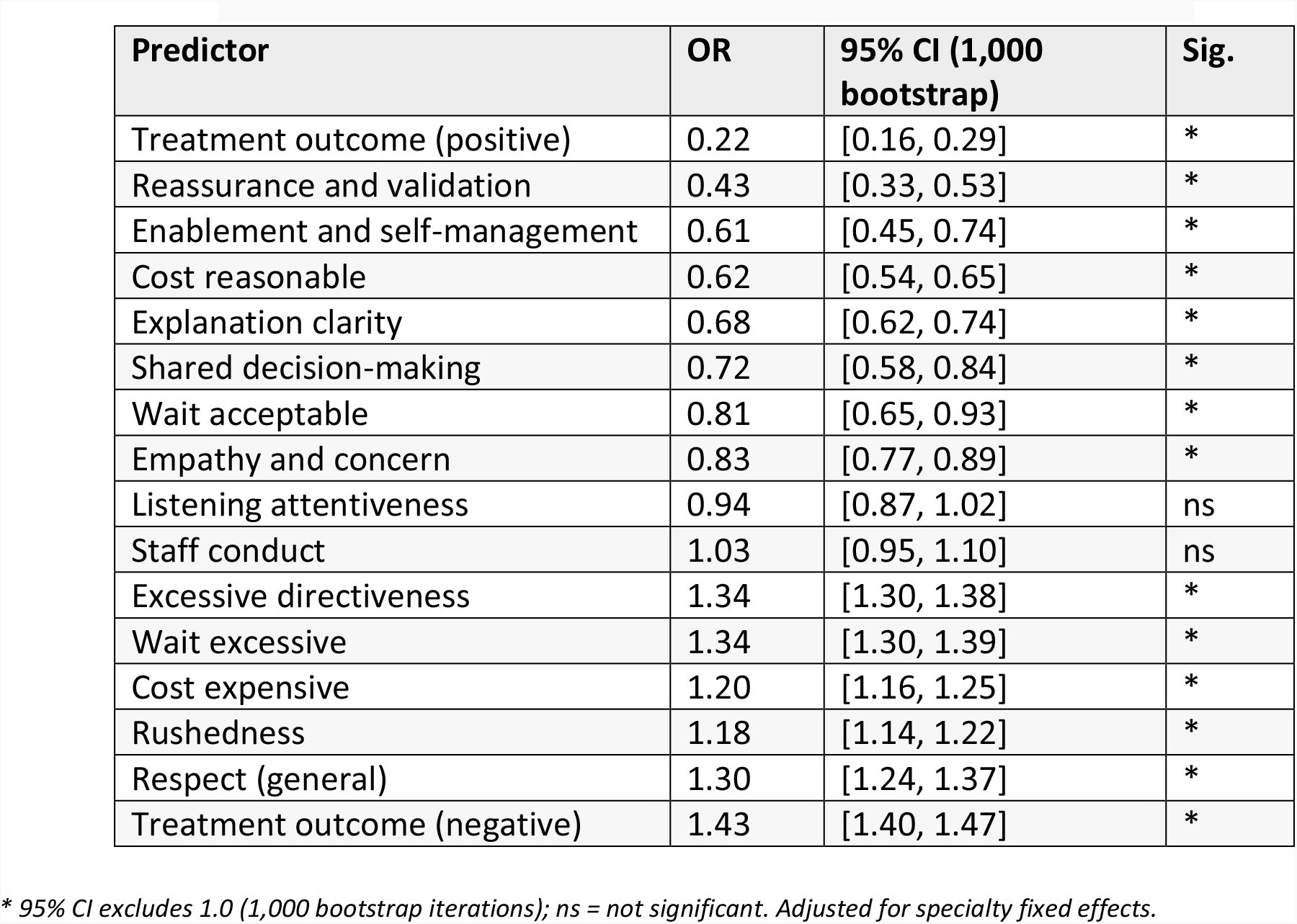
Logistic regression: switching intent (1=YES), n = 60,526.

**Figure 4.**
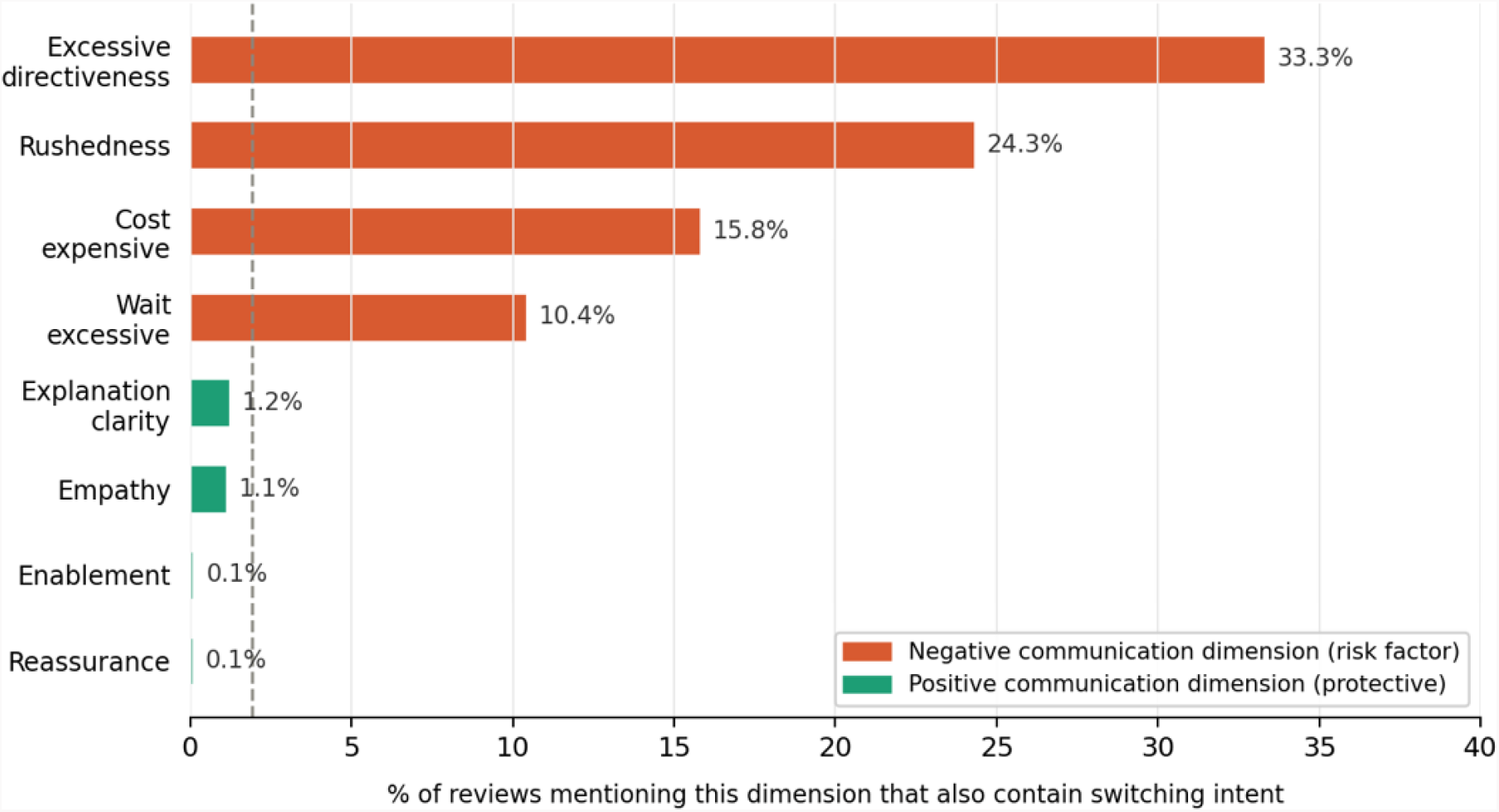
Switching intent rates by communication dimension. Bars show % of reviews mentioning each dimension that also contain switching intent language. Dashed line = overall switching rate (1.9%). n = 61,176.

## Discussion

This study applied the Kalamazoo Consensus Statement, Calgary-Cambridge Guide, and SPIKES protocol as a unified analytical framework to 61,176 patient reviews of Indian physicians across 10 cities and 8 specialties; the largest framework-anchored computational analysis of physician communication in the Indian context to our knowledge.

The principal finding is that communication quality is not a peripheral feature of patient experience but a central determinant of recommendation, retention, and defection. Two communication behaviors, excessive directiveness and rushedness, were associated with expressions of switching intent at rates 20-fold higher than in reviews where these behaviors were absent. Reassurance and empathy were the strongest positive predictors of recommendation. These effects were systematically modified by specialty in ways that align with the theoretical predictions embedded in the three frameworks.

### Asymmetric Effects of Communication Failures and Successes

Reassurance and empathy are the strongest positive predictors of recommendation, yet their absolute impact operates near a ceiling given the 93.8% positive base rate. Excessive directiveness and rushedness operate from a much lower floor, with unadjusted recommendation rates of 22.9% and 28.9% respectively when present. This asymmetric structure, where failures cost more than successes earn, is consistent with prospect theory ^[12]^ and the loss aversion literature. In practical terms, communication training programs targeting directive or time-pressured behaviors may yield greater returns on patient retention than those focused solely on amplifying warmth.

These asymmetric effects carry implications beyond patient satisfaction. In specialties where continuity of care is critical; endocrinology, diabetology, cardiology, and oncology, communication failures that drive defection can disrupt follow-up, worsen medication adherence, and delay escalation of care. Framing excessive directiveness and rushedness as modifiable risk factors for loss of continuity repositions communication training as a patient safety intervention rather than a purely experiential initiative.

### Specialty Context and the Directiveness Anomaly

Specialty context modifies which communication dimensions carry the most weight. Affective and validating behaviors, empathy and reassurance, are most prominent in psychiatry and oncology, where the patient-clinician relationship is itself part of the therapeutic process. Informational behaviors carry more relative weight in surgery and chronic disease management. The consistency between these observed specialty-specific patterns and theoretical expectations provides supportive evidence for the construct validity of the integrated coding framework.

Excessive directiveness shows a positive association with recommendation intensity across most specialties, yet is consistently the strongest negative predictor in the binary recommendation model. This apparent contradiction reflects how recommendation intensity was coded: intensity 3 captures emphatic signals in both positive and negative valence, and reviews mentioning directiveness are disproportionately emphatic regardless of direction. The binary model, which cleanly separates valence, shows the true effect. This anomaly is a property of the intensity coding design, not evidence of any clinical benefit from directive behavior.

### Cost Asymmetry in the Indian Context

The 9:1 ratio of negative to positive cost effects reflects India’s predominantly out-of-pocket healthcare financing model. Patients directly experience the price of consultations without insurance mediation, making pricing transparency a communication-adjacent concern with direct behavioral consequences. Physician practices and clinic operators in Indian private healthcare should treat fee communication as part of the consultation curriculum.

### AI-Assisted Analysis as a Tool for Communication Research

Beyond identifying communication behaviors associated with patient outcomes, a methodological contribution of this study is demonstrating that large language models can reliably apply established clinical communication frameworks to large-scale patient narratives. This enables scalable, theory-driven annotation that would be impractical using conventional manual coding, and provides a reproducible approach for communication research with applications in health services research, implementation science, and medical education. The approach is not offered as a replacement for observational or qualitative methods, but as a complement that makes large-scale hypothesis testing feasible within a theoretically grounded codebook.

### Implications for Medical Education, Clinical Practice, and Telehealth

The consistency of these findings across 61,000 reviews and eight specialties suggests that communication quality is not incidental to patient experience in Indian healthcare, it is central to it. Yet clinical communication receives minimal formal attention in Indian medical education.

The Medical Council of India’s MBBS curriculum introduced communication skills as a competency in 2019, but implementation remains uneven, assessment is largely formative, and specialty-specific communication training is essentially absent at the postgraduate level. The present findings suggest this gap has measurable behavioral consequences: patients whose physicians are perceived as directive or time-pressured show higher rates of switching intent; patients whose physicians are perceived as reassuring and attentive show higher rates of recommendation and retention. If communication behaviors are this strongly associated with patient retention and defection, the case for structured, specialty-specific communication training, assessed against patient-reported outcomes rather than faculty observation alone, is not merely pedagogical. It is commercially and institutionally consequential for Indian healthcare providers.

For postgraduate training programs, these specialty-stratified patterns offer a direct template for competency-based communication curricula. Psychiatry and oncology modules could prioritize reassurance and empathy in the context of chronic therapeutic relationships and bad-news delivery; surgical and procedural specialties could emphasize reducing directiveness and rushedness in pre-operative counselling and consent encounters; and chronic-disease specialties such as endocrinology and diabetology could systematically integrate enablement and self-management coaching into follow-up visit templates. Embedding such competencies within existing CBME structures, linked to workplace-based assessments and patient-reported outcomes, would allow communication training to be both specialty-specific and accountable.

For practicing physicians, the results reframe communication quality as an economic concern: communication behaviors associated with expressions of switching intent occurred at rates 20-fold higher in directive encounters than in non-directive ones. These behaviors are modifiable; the specialty-specific patterns identified here provide a basis for targeted, evidence-grounded feedback rather than generic communication audits.

For telehealth, now a substantial proportion of outpatient consultations in India, the implications may be sharper still. Digital encounters strip out the non-verbal and environmental cues that buffer poor communication in face-to-face settings, leaving verbal behavior as nearly the sole determinant of patient experience. The communication dimensions this corpus identifies as most damaging, directiveness and rushedness, are likely to carry even greater weight in video-mediated encounters where compensatory warmth cues are absent.

Because telehealth encounters are often recorded or documented in detail, they also provide a pragmatic substrate for structured communication audits and feedback cycles, enabling physicians to review concrete examples of directive or rushed behaviors and iteratively adjust their practice.

### Implications for Institutional Governance

At the institutional level, aggregated communication profiles by specialty and clinic could be incorporated into routine quality dashboards alongside traditional metrics such as waiting time, cancellation rates, and readmissions. Because excessive directiveness and rushedness show strong associations with switching intent and anti-recommendation, these dimensions are natural candidates for key performance indicators in hospital-level patient experience governance. Accreditation bodies and payers that increasingly incorporate patient feedback into evaluation frameworks may also benefit from communication-dimension metrics derived from large review platforms.

### Comparison with Prior Work

Dhakate and Joshi ^[2]^ identified broad positive and negative sentiment categories without framework anchoring or behavioral outcome analysis. The present study extends their work with a 1.6-fold larger sample, five-fold greater geographic coverage, framework-anchored dimension coding, and hypothesis-testing around recommendation, switching, and retention outcomes. Zhang et al.’s HSQ-5D ^[6]^ overlaps partially with our empathy and reassurance dimensions via their humanistic care construct, but clinical communication frameworks embed these as relational competencies with specific behavioral indicators rather than service quality attributes.

Unlike previous computational analyses of physician reviews, which have primarily relied on sentiment polarity or broad healthcare service-quality dimensions, the present framework operationalizes theoretically grounded communication behaviors derived from the Kalamazoo Consensus Statement, the Calgary-Cambridge Guide, and the SPIKES protocol. Rather than classifying reviews simply as positive or negative, it identifies specific communication behaviors that are directly interpretable within established models of clinical communication. This enables findings to be translated into competency-based medical education, structured communication audit, quality improvement initiatives, and implementation interventions in ways that generic sentiment or service-quality measures cannot readily support.

### Limitations

The corpus reflects patients who use Practo and write reviews, a self-selected urban, educated, smartphone-literate population. Rural and informal-sector patients are absent. The corpus contains no reviews from northeastern India, which has distinct healthcare infrastructure and patient population characteristics; findings should not be generalized to this region. Data quality concerns documented for physician-rating platforms ^[7]^ apply here, including positivity bias (93.8% positive) and potential for non-patient contributions despite screening exclusions. Bangalore over-representation (19.3%) and Kolkata under-representation (4.6%) mean specialty-city cell estimates are unequally reliable.

The coding procedure used expert confirmatory review of model output rather than fully independent double-coding. While 98.5% agreement across 200 reviews provides reasonable evidence of label consistency, the visibility of model predictions during review introduces the possibility of confirmation bias. Independent annotation on a fresh subsample would provide stronger reliability evidence and is a planned extension.

The analytical design is observational. Associations between communication dimensions and outcomes cannot be interpreted causally; unmeasured physician characteristics, patient case mix, and clinic-level factors confound the relationships observed. Communication dimensions were inferred from patient narratives rather than directly observed consultations and therefore reflect patient perceptions of physician behavior rather than objective measures of communication practice. Interventions targeting directiveness may improve retention and warrant prospective evaluation.

## Conclusions

This study applied three established clinical communication frameworks as a unified analytical scheme to 61,176 patient reviews of Indian physicians, identifying which communication dimensions predict recommendation, switching intent, and retention. Excessive directiveness and rushedness were the communication behaviors most strongly associated with adverse behavioral outcomes in this corpus; reassurance and empathy were the strongest drivers of recommendation. These effects vary by specialty in ways that align with the theoretical predictions embedded in the Kalamazoo, Calgary-Cambridge, and SPIKES frameworks, supporting the construct validity of the coding approach and the contextual specificity of communication expectations in Indian clinical practice.

Communication competence is unlikely to represent a single transferable skill across specialties: empathy is the universal currency, but reassurance carries particular weight in oncology and psychiatry, explanation and shared decision-making in surgery, and enablement in endocrinology and diabetology.

For physician communication training, the findings suggest that reducing directive and time-pressured consultation behavior may yield greater impact on patient retention than amplifying warmth, though both matter significantly.

Future research should validate this framework across other healthcare settings, languages, and physician-rating platforms, and examine whether similar communication patterns are observed in directly recorded clinical encounters and teleconsultations. Although the present study did not evaluate teleconsultations specifically, the identified communication behaviors are likely to remain relevant in digitally mediated consultations, where verbal interaction constitutes a greater proportion of the patient experience. Prospective studies evaluating interventions designed to improve high-impact communication behaviors will further clarify how theory-informed communication training translates into measurable improvements in patient-centered outcomes.

## ACKNOWLEDGEMENTS

The authors thank the patients whose publicly available reviews form the dataset for this study. We acknowledge the use of GPT-4.1-mini (OpenAI) for LLM-assisted annotation and Claude Sonnet (Anthropic) for pilot coding.

## AUTHOR CONTRIBUTIONS (CRediT taxonomy)

RM: Conceptualization, Methodology, Data curation, Formal analysis, Investigation, Validation, Visualization, Writing – original draft, Writing – review and editing.

SS: Conceptualization, Project administration, Supervision, Writing – review and editing.

BK: Conceptualization, Methodology, Writing – review and editing, with specific contribution to clinical framing of medical education and institutional governance implications.

MM: Conceptualization, Methodology, Writing – review and editing, with specific contribution to clinical framing of medical education and institutional governance implications.

VE: Conceptualization, Methodology, Writing – review and editing, with specific contribution to clinical framing of medical education and institutional governance implications.

## CONFLICT OF INTEREST STATEMENT

RM and SS are employees of SigmaMozak Solutions Pvt. Ltd., which develops ConversationAIly−, an AI-based clinical communication training product. The analytical framework and dataset used in this study are entirely independent of ConversationAIly−; the study does not use, evaluate, or commercially promote that product. The authors declare no other conflicts of interest. BK, MM and VE have no conflicts of interest.

## FUNDING STATEMENT

This research received no external funding. Data collection, annotation, and analysis were conducted using internal resources of SigmaMozak Solutions Pvt. Ltd. LLM annotation costs were approximately USD 100.

## ETHICS STATEMENT

This study used publicly available patient review data collected from publicly accessible physician profile pages on the Practo platform. No login, authentication, subscription, or bypass of technical access controls was required. No individually identifiable patient information was collected or processed. Physician identities were pseudonymized using platform-assigned slugs. The study analyzed existing publicly available content without interacting with users or altering platform functionality. No ethics committee approval was required under applicable guidelines for secondary analysis of publicly accessible digital-trace data. No informed consent was required.

## DATA AVAILABILITY STATEMENT

The codebook, annotation pipeline, and statistical analysis scripts are publicly available at https://doi.org/10.17605/OSF.IO/UARJW. The full labeled dataset is available from the corresponding author on reasonable request. Original patient reviews remain publicly accessible at their source URLs on practo.com. The dataset is not deposited in a public repository because it contains physician names and clinic details that, while individually public, could facilitate identification of practitioners when aggregated.

## Supplementary Materials

**Table A1.** Expert confirmatory review: label-level agreement (n = 200 stratified reviews)

| Dimension | Cohen's k | Agreement % | Reviewer % | GPT % | Changed |
| --- | --- | --- | --- | --- | --- |
| Empathy and concern | 0.940 | 97.0% | 46.0% | 46.0% | 6 |
| Listening attentiveness | 0.965 | 98.5% | 30.0% | 31.5% | 3 |
| Explanation clarity | 0.978 | 99.0% | 36.0% | 37.0% | 2 |
| Enablement | 0.960 | 99.5% | 7.0% | 6.5% | 1 |
| Respect (general) | 0.898 | 98.0% | 10.5% | 11.5% | 4 |
| Shared decision-making | 0.945 | 99.5% | 5.0% | 4.5% | 1 |
| Excessive directiveness | 1.000 | 100.0% | 3.0% | 3.0% | 0 |
| Rushedness | 1.000 | 100.0% | 3.5% | 3.5% | 0 |
| Reassurance | 1.000 | 100.0% | 21.5% | 21.5% | 0 |
| Staff conduct | 0.981 | 99.5% | 15.5% | 15.0% | 1 |
| Switching intent | 0.808 | 98.0% | 6.5% | 4.5% | 4 |
| Mean / Total | 0.952 | 98.5% | — | — | 22 |

Reviewer: lead author (RM). GPT-4.1-mini predictions visible during review; labels confirmed or overridden. Expert confirmatory review of model output, not fully independent double-coding. Cohen's kappa reported for completeness.

**Table A2.** Inter-model calibration: GPT-4.1-mini vs. Claude Sonnet (n = 200 pilot reviews)

| Dimension | Cohen's k | Agreement % | Verdict |
| --- | --- | --- | --- |
| Listening attentiveness | 0.924 | 96.5% | Pass |
| Explanation clarity | 0.943 | 97.5% | Pass |
| Excessive directiveness | 0.828 | 99.0% | Pass |
| Reassurance | 0.804 | 96.0% | Pass |
| Staff conduct | 0.914 | 98.0% | Pass |
| Rushedness | 1.000 | 100.0% | Pass |
| Empathy | 0.789 | 89.5% | Pass |
| Rec. intensity | — | 47.0% | Scale compression, binary outcome designated primary (see Section 2.4) |

Threshold: $k \geq 0.60$ = pass. Expert confirmatory review confirms GPT-4.1-mini recommendation intensity mean (1.45 reviewer vs. 1.47 model; 99.5% exact agreement).

## Notes

### Competing Interest Statement

The authors have declared no competing interest.

## References

[1] Hong YA, Liang C, Radcliff TA, Wigfall LT, Street RL. What do patients say about doctors online? A systematic review of studies on patient online reviews. J Med Internet Res. 2019;21(4):e12521. doi:10.2196/12521

[2] Dhakate N, Joshi R. Classification of reviews of e-healthcare services to improve patient satisfaction: insights from an emerging economy. J Bus Res. 2023;164:114015. doi:10.1016/j.jbusres.2023.114015

[3] Makoul G. Essential elements of communication in medical encounters: the Kalamazoo consensus statement. Acad Med. 2001;76(4):390–393. doi:10.1097/00001888-200104000-00021

[4] Kurtz SM, Silverman JD. The Calgary-Cambridge Referenced Observation Guides: an aid to defining the curriculum and organizing the teaching in communication training programmes. Med Educ. 1996;30(2):83–89. doi:10.1111/j.1365-2923.1996.tb00724.x

[5] Baile WF, Buckman R, Lenzi R, Glober G, Beale EA, Kudelka AP. SPIKES—a six-step protocol for delivering bad news: application to the patient with cancer. Oncologist. 2000;5(4):302–311. doi:10.1634/theoncologist.5-4-302

[6] Zhang X, Sun J, Li X, Liu Y, Li C. Developing a framework for online review-based health care service quality assessment: text-mining study. J Med Internet Res. 2025;27:e66141. doi:10.2196/66141

[7] Mulgund P, Sharman R, Anand P, Shekhar S, Karadi P. Data quality issues with physician-rating websites: systematic review. J Med Internet Res. 2020;22(9):e15916. doi:10.2196/15916

[8] Hao H. The development of online doctor reviews in China: an analysis of the largest online doctor review website in China. J Med Internet Res. 2015;17(6):e134. doi:10.2196/jmir.4365

[9] Gilardi F, Alizadeh M, Kubli M. ChatGPT outperforms crowd workers for text-annotation tasks. Proc Natl Acad Sci USA. 2023;120(30):e2305016120. doi:10.1073/pnas.2305016120

[10] Anthropic. Claude model card: Claude Sonnet. Anthropic; 2025. https://www.anthropic.com/research/model-card

[11] OpenAI. GPT-4.1 system card. OpenAI; 2025. https://openai.com/index/gpt-4-1

[12] Kahneman D, Tversky A. Prospect theory: an analysis of decision under risk. Econometrica. 1979;47(2):263–291. doi:10.2307/1914185

